# A within-person randomised trial to investigate the effects of a rigid cervical collar on three-dimensional angular movement and angular acceleration during emergency spinal immobilisation and extrication procedures in elite football (soccer) players: The RESTRICT study

**DOI:** 10.1101/2023.11.22.23298822

**Authors:** Michael J Callaghan, Tom Hughes, John Davin, Russell Hayes, Neil Hough, Dan Torpey, David Perry, Sam Dawson, Eoghan Murray, Matthew Lancett, Anmin Liu, Richard K Jones

## Abstract

**Objective:** To determine the effects of rigid collars on cervical movement and acceleration during triple spinal immobilisation and extrication.

**Methods:** Procedures were performed on 15 non-injured volunteers in random order of collar and no-collar. Primary outcomes were angular movements and angular accelerations of the head relative to the thorax. Secondary outcome was the total procedure time.

**Results:** Between collar and no-collar, small but significant differences were found for mean angular movements during 15^°^ tilt tasks for lateral flexion (3.4^°^: 95%CI: 1.4^°^, 5.4^°^), flexion-extension (2.4^°^ 95%CI: 0.4^°^, 4.4^°^), rotation (–3.7^°^: 95%CI: –7.2^°^, –0.2^°^) and total rotation (6^°^: 95%CI; 2.9^°^, 9.1^°^). For the lift and lower task there was a significant difference in total lateral flexion of only 0.3^°^ (95%CI: 0.1^°^, 0.5^°^). Total movement during the procedure was significantly more for rotation without the collar (6.6^°^: 95% CI: 1.9^°^, 11.3^°^). Small but significant differences were found for angular acceleration only during the lift and lower task for maximum lateral flexion (–6.5 rads/s^2^, 95%CI: –12, –1 rads/s^2^), maximum rotation (–2.8 rads/s^2^, 95% CI: –5.2, –0.4 rads/s^2^) and minimum rotation (–3.5 rads/s^2^, 95%CI: –5.9, –0.1 rads/s^2^). The procedure was significantly longer with the collar (257.5s [95%CI: 245.3, 269.7s} *versus* 230.9s [95%CI: 215, 246.8s].

**Conclusion:** There were statistically significant but clinically negligible differences between a rigid cervical collar and no-collar in some parameters for the triple immobilisation and extrication procedure in the sporting context. These novel results provide highlight important clinical considerations when immobilising and extricating players after a head or cervical injury.

## INTRODUCTION

Major traumatic cervical spine injuries in sport are rare, yet can have potentially devastating sequelae, such as spinal cord injury (SCI) with associated neurological impairment and premature mortality^1, 2^. If an athlete suffers loss of consciousness, or a destabilising cervical injury cannot be ruled out, the whole spine should be immobilised to reduce the likelihood of further or secondary SCI injury due to hypoperfusion and hypoxia^1^.

The most effective methods of cervical spinal immobilisation are unclear and controversial,^3^ but typically include applying a rigid cervical collar and securing the individual to a stretcher with body straps, head blocks and tape or straps^4^ (so called ‘triple immobilisation’).

In triple immobilisation, rigid collars may independently safeguard the injured cervical spine from adverse motion to a limited extent^5^ and are recommended in many pre-hospital care guidelines^1, 2, 5–7^ as well as advanced trauma courses for sport in the United Kingdom^8^. Despite their widespread use, recent Danish guidelines suggest that the use of collars should be completely avoided, although this recommendation is based upon weak evidence^9^. This reflects the consensus statement from the Faculty of Pre-Hospital Care highlighting the growing concerns of using collars^4^.

These different recommendations may be explained because there are few studies supporting the beneficial effects of rigid collars on neurological and survival outcomes, compared to the mounting evidence of adverse effects^9^, such as airway compromise, increased intracranial pressure and patient distress^5^. Furthermore, a Cochrane review^10^ suggested that discomfort from cervical immobilisation might result in conscious individuals repositioning themselves, which could theoretically worsen any existing spinal injuries.

The general sub-optimal quality of the existing evidence has also made it difficult to establish the independent efficacy of rigid cervical collars as part of the immobilisation procedure^5, 11^. Cadaveric studies^12, 13^ have limited external validity. Some human studies using a collar with spinal boards^11, 14, 15^ or head blocks^16^ analysed immediate collar application rather than the whole triple immobilisation and extrication procedure. Dixon et al.^17, 18^ used infra-red motion analysis of immobilisation and extrication from a seated position in simulated road traffic accidents. Up to 5.5^°^ more cervical spine movement was recorded during extrication <u>with</u> a collar and usual pre-hospital rescue equipment than controlled no-collar self-extrication. However, these data are unlikely to generalise to sports trauma scenarios. To date there are no known studies that have investigated the effects of collar *versus* no-collar conditions on the cervical spine kinematics during immobilisation and extrication methods in sports trauma management. Moreover, a systematic review^19^ exposed the existing gaps in our basic knowledge regarding cervical spine external stabilisation and exhorted researchers to investigate further the effects of cervical immobilisers.

Therefore, our aim was to measure three-dimensional angular movement and angular acceleration of the head relative to the thorax during a full spinal triple immobilisation and extraction procedure from a football (soccer) pitch, with and without a rigid cervical collar. We hypothesised that tasks performed with a rigid cervical collar would have significantly less angular movement and acceleration and take significantly longer to complete than tasks without a collar.

## METHODS

### Design

The Range of movement Evaluation using Stabilisation Techniques during extRaction In Cervical Trauma (RESTRICT) study was a prospective, within person randomised design trial under two conditions: namely, the current practice in sport of immobilisation and extrication *with* and *without* a cervical collar. Each participant served as their own control.

The study was conducted and reported according to the recommendations of the Consolidated Standards of Reporting Trials (CONSORT) statement extension for within person randomised trials^20^. A detailed protocol of the RESTRICT study has been published elsewhere^21^.

There were two deviations from the protocol. Firstly, due to the availability of injury free players, the age range was altered to 16 – 21 years (from 18 – 23 years). Secondly, the primary outcome was captured and recorded by the Xsens inertial-sensor-based motion capture system (Xsens Technologies, XSens MTx, www.xsens.com) instead of the Delsys system. Data were collected between May 2022 and August 2022, on a 4^th^ generation AstroTurf™ pitch at an indoor training facility at an English Premier League Football Club.

There was no external funding. The study was registered before active recruitment commenced (<u>International Standard Randomised Controlled Trial Number (ISRCTN) Registry:</u> https://doi.org/10.1186/ISRCTN16515969) and was ethically approved by the University of Salford (reference number 1403).

### Participants

Participants who simulated the role of players with a head or neck injury were a convenience sample from a cohort of non-injured, elite football players under contract at one English Premier League Football Club. Participants were eligible for inclusion if they were between 16 and 21 years old at the time of the study, but were ineligible if they were being treated for any musculoskeletal injury or illness. All participants provided informed written consent.

There was staff pool of nine physiotherapists and two medical doctors. Six clinical practitioners from the pool performed each procedure. They were English Football Association (FA) level 5 Advanced Trauma and Medical Management in Football (ATMMiF) trained members of medical staff at the same English Premier League Football Club. Neither the clinical practitioners nor the participants were blinded to the randomised conditions.

### Procedure

The procedure consisted of full spinal triple immobilisation according to current ATMMiF course guidelines^8^. The procedure was divided into five tasks. Each was considered a key component likely to affect cervical angular movement and angular acceleration (Table 1; figure 1). The randomisation, procedure and equipment are fully described in the protocol^21^. Briefly, each procedure was performed with the collar given sequentially, (three times *with* and three times *without)* without removing the IMUs, using the Laerdal Stifneck Select Cervical collar (Laerdal Medical, Stavanger, Norway).

**Figure 1:** Illustrates the sequence for the triple spinal immobilisation and extrication procedure MedRxiv policy is to avoid the inclusion of photographs and any other identifying information of people, whether it be authors, patients, participants, test volunteers or experimental stimuli, because verification of consent is incompatible with the rapid nature of preprint posting. Therefore, readers may contact the corresponding author to request access to these images.

**Table 1.**
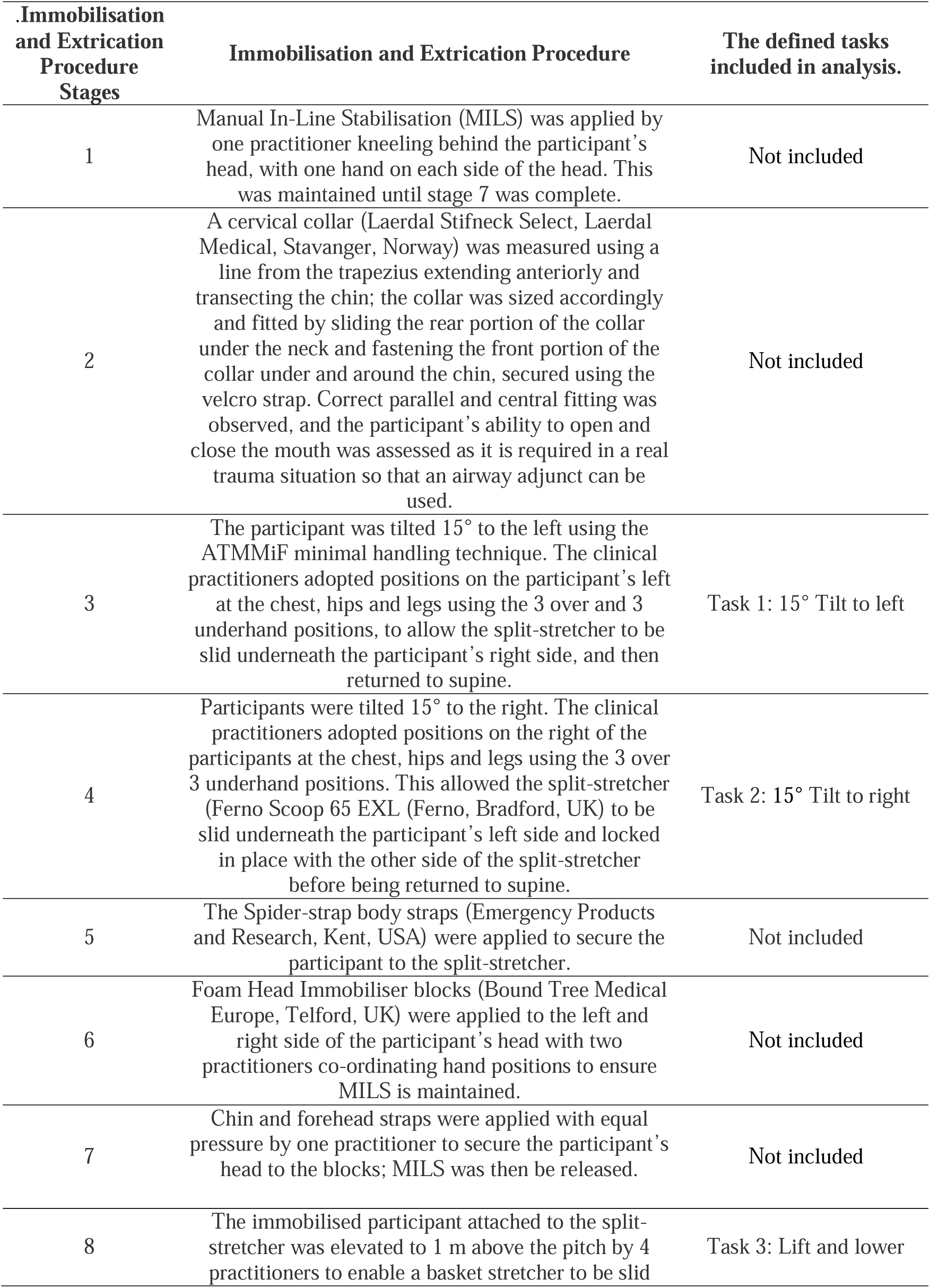
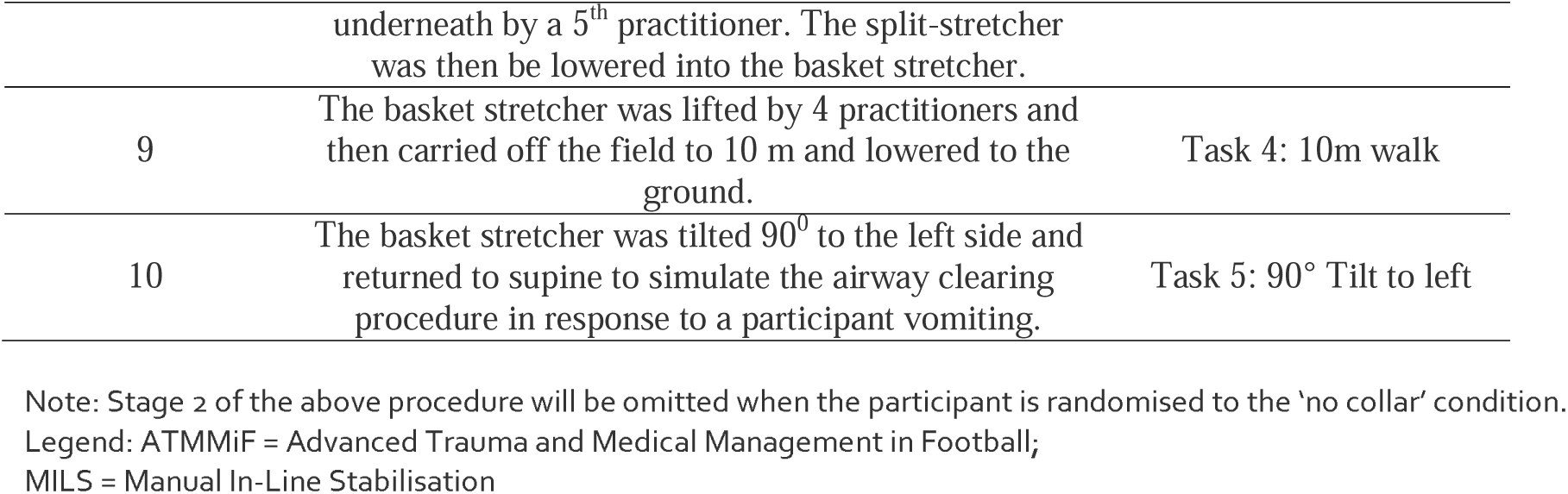
Description and identification of procedural stages for the whole procedure and the defined task labels which are included in the analysis.

### Outcome measures

The primary outcomes were the differences between collar and no-collar conditions for 1) mean and total angular movement (i.e. range of movement – degrees) and 2) minimum and maximum angular acceleration (i.e. speed of movement – radians per second^2^) of the head relative to the trunk from an initial neutral position for flexion, extension, rotation, and lateral flexion during each task. The secondary outcome was the mean time taken to fully complete each immobilisation procedure from the beginning of task 1 to the end of task 5.

### Sample Size

An *a priori* sample size calculation was performed from acceleration data presented in McDonald et al.^22^ With a mean difference between patients of 2.8 m/s^2^ (95% CI: 10.6=1 SD±3.42), a within-subjects effect size of 0.5, with power set at 0.8 (the probability of a type-II error) and an alpha level of 0.05 (the probability of a type-I error) the number of player participants required was n=15.

## DATA CAPTURE

The Xsens inertial sensor based motion capture system has excellent intra-rater reliability for all cervical motion metrics in the axial plane: ICC values of 0.94 (95% CI: 0.90–0.97) for axial flexibility; 0.94 (95% CI: 0.89–1.0) for velocity; 0.92 (95% CI: 0.83– 1.0) for acceleration measures^23^. Nine sensors were secured to the head on top, chest on sternum, both shoulders, pelvis and lower and upper arms of both sides with neoprene proprietary attachments. Data were samples at 100Hz, transferred wirelessly, saved to a laptop computer, and checked for signal loss or artefact. The data for each task were manually time-stamped by a researcher (AL) observing the procedure.

## STATISTICAL ANALYSES

For all parameters, mean point estimates and corresponding 95% confidence intervals (CIs) were calculated for each participants’ three trials with and three trials without the collar. The analysis was based on paired data. For angular movement, mean and total movement values for three trials in all planes are presented in degrees. For angular acceleration, minimum and maximum values are presented in radians/second^2^ (Tables 2 and 3). For the secondary outcome of time taken to complete the whole procedure, mean point estimates and 95%CIs were compared between conditions. All data were explored for normality using the Shapiro-Wilk test. For all parameters, the estimated mean within-subject differences between each condition (collar *versus* no-collar) were calculated for each of the five tasks and across the whole procedure with corresponding 95% CIs (Excel, Microsoft Inc USA). The statistical analyses and presentation are consistent with the Checklist for statistical Assessment of Medical Papers (CHAMP) statement^24^.

**Table 2:**
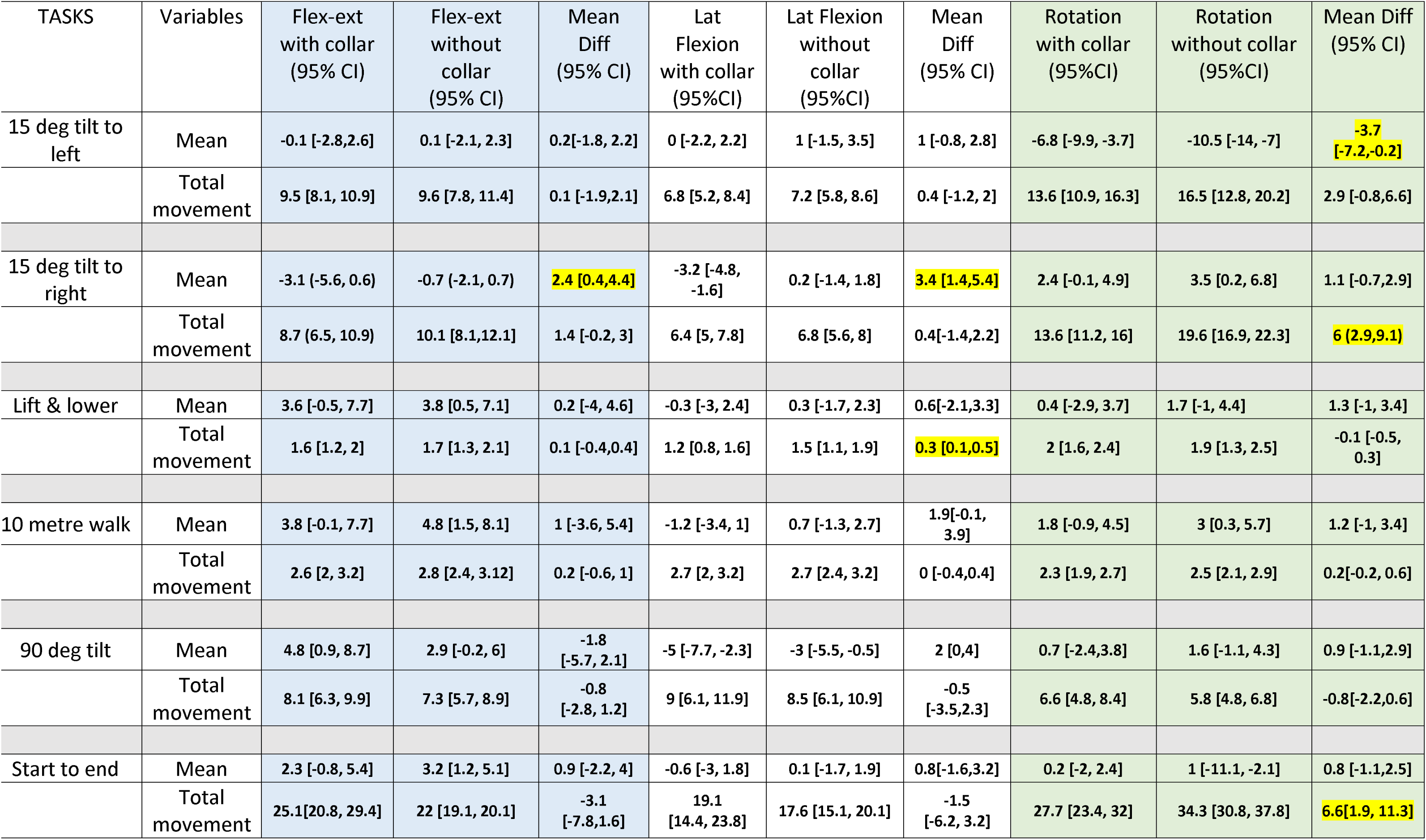
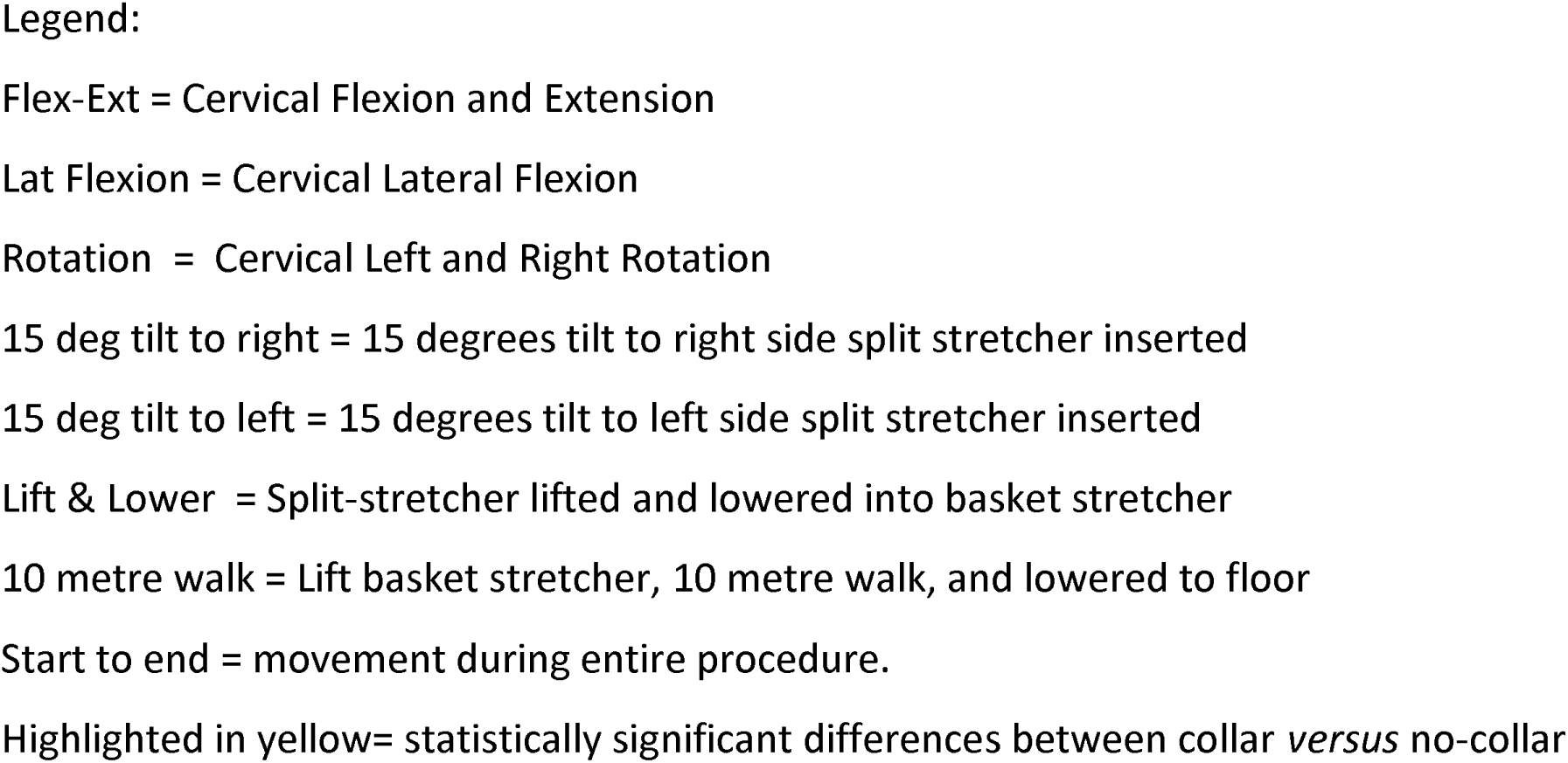
The angle (degrees, 95% CI) of the head relative to the thorax during different tasks; 3 trials with and 3 trials without collar.

**Table 3:**
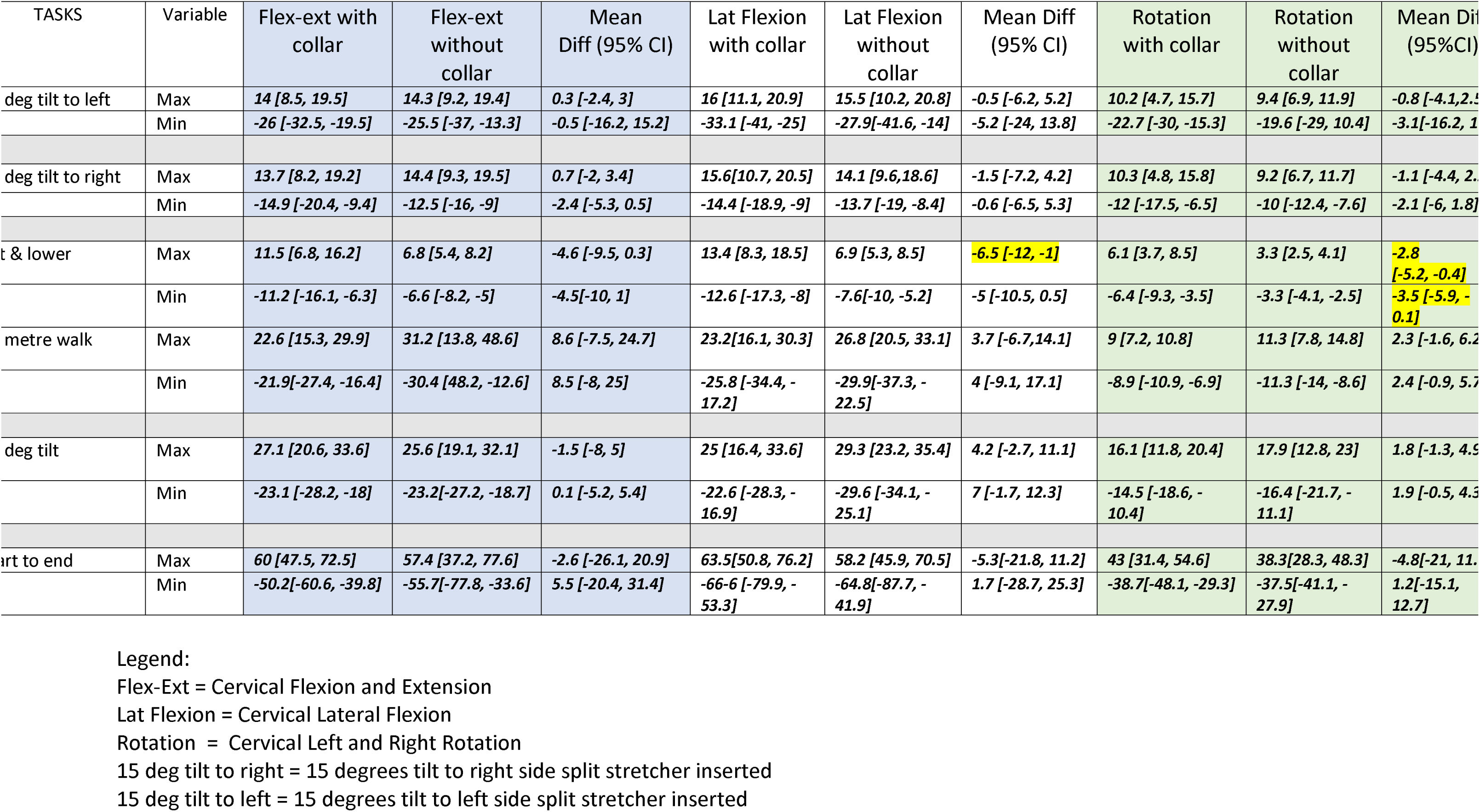
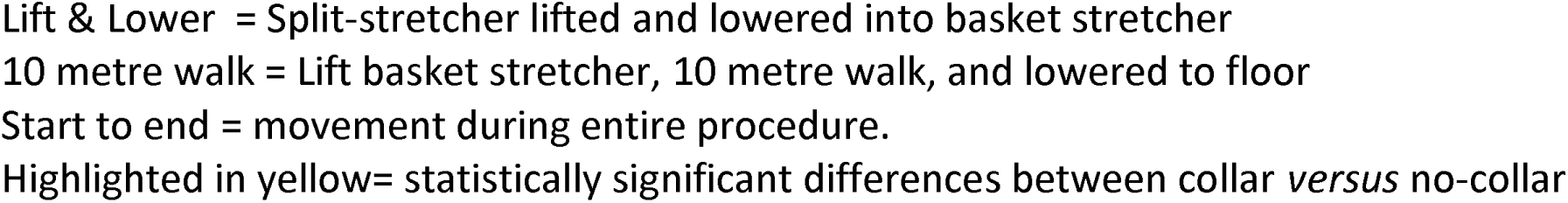
The angular acceleration (rads/s^2^, 95%CI) of the head relative to the thorax during different tasks 3 trial with and 3 trials without collar.

## RESULTS

Fifteen male participants aged 17.7 years (± 1.4), body mass 71.1kg (±8.1), height 1.70m (±0.01) and body mass index 24.6kg/m^2^ (±2.8) volunteered. One participant’s data were excluded in the final analysis due to data capture errors. All remaining data were complete. No adverse effects were reported by participants or clinicians. All recorded data were normally distributed.

### Angular movement

Across the whole procedure (task 1 to task 5), only total rotation was significantly restricted with a collar but with wide 95% CIs (mean difference = 6.6^°^; 95%CI: 1.9°, 11.3°) (Table 2). In only two tasks, the *total* angular movement was significantly more restricted with the collar and only by a neglible amount. Firstly, the collar restricted rotation during the 15^°^ right tilt task, (mean difference = 6^°^ (95% CI: 2.9°, 9.1°), and for lateral flexion during the lift and lower task (mean difference = 0.3^°^; 95%CI: 0.1°, 0.5°) (Table 2).

The *mean* angular movement was significantly more restricted with the collar in only two tasks. The 15^°^ right tilt task had a small but precisely estimated differences in lateral flexion (mean difference = 3.4^°^: 95%CI; 1.4°, 5.4°) and flexion-extension (mean difference = 2.4^°^: 95%CI: 0.4°, 4.4°). The 15^°^ left tilt task had a small difference in rotation, but with wider CIs (mean difference = –3.7^°^: 95%CI: –7.2°, –0.2°) (Table 2).

### Angular acceleration

For angular acceleration (speed of movement), a statistically significant difference between collar and no-collar conditions was observed only during the lift and lower task. Notably, collar application resulted in marginally increased but negligible acceleration values for maximum lateral flexion (6.5 rads/s^2^; 95%CI: –12, –1 rads/s^2^; maximum rotation –2.8 rads/s^2^; 95% CI –5.2, –0.4 rads/s^2^ and minimum rotation –3.5 rads/s^2^; 95%CI: –5.9, –0.1 rads/s^2^); the 95% CIs indicated that these estimates were precise (see Table 3).

### Time taken to complete procedure

The mean time taken for the whole procedure was significantly longer with the collar 257.5s [95%CI: 245.3, 269.7 secs] *versus* 230.9s [95%CI: 215, 246.8s without.

## DISCUSSION

This study is the first to examine the effects of rigid collar application on cervical movement and angular acceleration profiles in football (soccer) players during a simulated spinal immobilisation and extrication procedure, as recommended in the FA ATMMiF course and many pre-hospital care guidelines^1, 2, 5–7^. Overall, we found that only 6.25% of parameters (nine out of 144) had statistically significant differences between the collar and no-collar conditions.

### Angular movement

Our first hypothesis was only partially proven. Across the whole procedure (i.e. task 1 to task 5), collar application offered a 6.6^°^ restriction in total rotation (albeit with wide 95% CIs). However, this was likely driven by the significant differences between conditions for the 15^°^ tilt tasks (tasks 1 and 2). For the other parameters, using a collar offered no superiority for restriction of cervical movement (apart from the significant but neglible mean difference in lateral flexion during the lift and lower task). Differences observed during 15^°^ tilt tasks were very small for mean movement (range 2.4° to 3.7°) and total movement (range 0.1° to 6°). Assessing the clinical significance of these differences is difficult as the movement threshold for causing or worsening neurologic injury is unknown. There are no comparable angular movement data for our specific immobilisation and extrication procedure in the sporting context, so our study provides novel insights.

We used an aggressive 90^°^ tilt task to simulate an emergency counter measure for players vomiting and found no significant differences between the collar and no-collar condition for all movement planes. These results should only be compared with caution to a 90° log roll with MILS and collar^25–27^, as our 90^0^ tilt task was performed with tape, head blocks and body straps, with a split stretcher and loaded into a basket stretcher. Our mean movement results are below the minimally important differences of 3^°^ for lateral flexion and rotation and 5^°^ for flexion/extension^25–27^. The minimal movement in our 90^0^ tilt task may be explained in the systematic review by Holla et al^19^, who suggested that the level of immobilisation generally increased as the surface area support increased and external support of the cranium and thorax provided nearly complete immobilisation. Devices (or practitioners) that just support the head and cervical area can only restrict by half the normal range of movement. Additionally, our study confirms that once an athlete is fully immobilised with tape, head blocks, body straps, placed on a split stretcher and loaded into a basket, there is sufficient restriction of head movement without the need for a rigid cervical collar.

To extricate a player, guidelines suggest using a either split stretcher or a spinal board to lift into a basket stretcher, before extricating from the pitch. A split stretcher requires a right and left 15^°^ tilt, whereas a spinal board requires 90^0^ log roll. Our data show that 15^°^ tilt tasks had less range of movement in all planes than the four person 90^0^ log roll used by Shrier et al^26^. Their larger values compared with ours for collar and no-collar indicate that to insert an extrication device, a 15^°^ tilt manouvre with or without a collar minimises cervical movement compared to a 90^0^ log roll.

### Angular acceleration

Our study is the first to report angular acceleration (speed of movement) of the head during the triple immobilisation and extrication procedure in sport, so provides novel data. Linear accelerations may have a role in tissue damage in the pre-hospital setting, and have been reported in other studies^22,27,26^. However, linear accelerations are not as comprehensive as the angular accelerations used in our study, due to their sensitivity to sensor location. Our sensors were placed on top of the head, to ensure that rotation (about the Z axis) might be detected. Additionally, we have presented maximum and minimum acceleration values and not mean values (Tables 2 & 3) as this would represent clinically relevant changes in acceleration during the tasks. As individuals moved to and from a starting position, the mean value would not represent these changes. Clinically significant thresholds are difficult to establish for angular acceleration. The lift and lower task was the only time where using a collar resulted in statistically significant mean reductions in angular acceleration with negligible values for lateral flexion (6.5rad/s^2^; 95%CI: –12, –1) and rotation (2.8 rad/s^2^ 95%CI: –5.2, –0.4). Across the whole procedure, the maximum rotation was 17.9 rad/s^2^ (95% CI: 12.8, 23 rad/s^2^) during the 90° tilt task. In context, this is only 0.62% of 2877 rad/s^2^, which is the threshold of angular rotation acceleration needed to cause mild concussion without loss of consciousness, and only 0.22% of the 8000 rad/s^2^ needed for severe concussion^28^. This demonstrates that all head movements occurring during the procedure had extremely low angular acceleration values and very low potential for further injury.

### Time taken to complete the procedure

Perhaps unsurprisingly, our second hypothesis was proven, because the immobilisation and extrication procedure took on average nearly half a minute faster without applying a collar. While there is no guidance on the optimal time to complete a triple immobilisation and extrictation procedure, our data show that the additional time to correctly fit a cervical collar could result in a delay to initiation of specialist treatment in time-critical patients (such as those with a serious, developing head injury).

## IMPLICATIONS FOR PRACTICE

Using rigid cervical collars as part of the immobilisation and extrication process following cervical spine injury is controversial because of the limited evidence of their efficacy, effects on neurological and survival outcomes, and increasing evidence of adverse effects^9^. A consensus group^4^ strongly advocated change from a policy of complete immobilisation to a system of selective immobilisation designed to reduce the risks of further injury.

Our results support this approach as in most parameters using a collar offered no superiority restricting cervical movement and acceleration. During the 15^°^ tilt tasks we found while a collar offers greater restriction of movement, the negligible acceleration values suggest this may not be of clinical importance.

Overall, our data suggest that a collar may be applied during 15^°^ tilt tasks when inserting a split-stretcher extrication device and may be loosened this once the head blocks and straps are in situ and the player is placed into a basket stretcher. This might negate some of the adverse effects previously associated with collar application without compromising the restriction to angular movement and acceleration. However, we recommend that this is context specific. In cases which are time critical (e.g serious developing head injury) then clinicians should favour the speed of extrication over cervical collar application.

## LIMITATIONS and STRENGTHS

In terms of study strengths, the crossover design avoided some of known parallel trial disadvantages of larger dropout rate, confounding bias, variability, instability of the participant’s condition. Potential carry-over effects were mitigated by the intervention. All the practitioners performing the procedure were FA ATMMiF level 5 trained members thus ensuring a high level of procedural consistency, skill and expertise. The order of condition was randomised and adequate sample size was calculated.

The limitations were that neither the clinicians, players, nor those collecting the data could be blinded to the randomised conditions. Additionally, conscious, non-injured players without painful muscle spasm were used, so the potential for neck movement in this cohort was likely more than an injured conscious player which may limit generalisability. Although we recommend further research to confirm our findings and to investigate this in realistic scenarios, we acknowledge the potential medico-legal, ethical, and consent barriers to conducting such studies on an acutely injured player.

## CONCLUSION

Within the sporting context we have shown that there are statistically significant but clinically negligible effects of using rigid cervical collar in a very small number of angular movement and acceleration parameters during the triple immobilisation and extrication procedure on non-injured players. The time taken to complete the procedures without the collar was significantly less. These results provide new knowledge and highlight important clinical considerations when immobilising and extricating players who have sustained a head or cervical injury.

## AUTHORS’ STATEMENT AND CONTRIBUTORSHIP

MJC, TH and RKJ drafted the manuscript.

AML completed the biomechanical modelling and computations. Completed all the results tables.

MJC, TH, JD, NH, DP, RH, DT, EM, SD, participated in the design and preparation of the study.

JD, NH, DP, RH, DT, EM, SD, ML, MJC, TH, AML, RKJ executed the study and data collection.

MJC, TH, AML, RKJ critically revised the manuscript’s revisions. RKJ, AML, TH and MJC handled and analysed the data.

MJC, TH, JD, NH, DP, RH, DT, EM, SD, ML, AML and RKJ read and approved the final revised version of the manuscript.

## DATA SHARING STATEMENT

An anonymised summary of the dataset to be analysed during this study may be available from the corresponding author on reasonable request.

## Data Availability

All data produced in the present study are available upon reasonable request to the authors.

## ACKNOWLEDGEMENTS

We wish to sincerely thank Dr Steve McNally, formally Head of Medicine at Manchester United FC for his support for this study. We also gratefully acknowledge the statistical advice of Dr Jamie Sergeant at the University of Manchester. We also thank Mr Lumbani Muntahi.

